# Implementation of a mobile app (iMPAKT) for improving person-centredness in nursing and midwifery practice: Protocol for a multi-methods evaluation study

**DOI:** 10.1101/2024.05.02.24306741

**Authors:** Sean R O’Connor, Donna Brown, Ian Cleland, Valerie Wilson, Tanya V McCance

**Affiliations:** Institute of Nursing and Health Research, Ulster University; School of Computing, Ulster University; South East Sydney LHD & University of Wollongong

## Abstract

The iMPAKT App has been developed as a digital tool for implementing and measuring person-centredness in nursing and midwifery practice. Despite its potential usefulness for the collection of person-centred measures, appropriate strategies are required to enhance the implementation of the app. To better understand the factors affecting adoption and maintenance, this protocol describes a multi-methods study to examine the experience of using the iMPAKT App in different contexts and settings. A convergent, multiple-methods approach will be used. Nurses and midwifes working in teams at different study sites in the UK and Australia will use the app during two, six-week cycles of data collection. Qualitative interviews and focus groups, guided by the Consolidated Framework for Implementation Research (CFIR) will be used to explore individual responses, views and experiences around acceptability and engagement with the app, and to examine variations in contexts. Quantitative data will be gathered on the number of person-centred measures recorded during the data collection cycles and using the System Usability Scale. Results will help to develop an understanding of the determinants and processes underpinning successful implementation, and inform further research to develop tailored implementation strategies, aimed at facilitating large scale collection of data on person-centred measures using the iMPAKT App.

## INTRODUCTION

There is an increasing global commitment towards creating more person-centred healthcare systems. An expanding body of evidence has established a clear association between person-centred practice and improvement in quality outcomes[1]. Person-centred practice is described as an approach which is established through healthful relationships between care providers, service users and others who are significant to them, including family members or informal carers[2]. Underpinning values include respect for the persons individual right to self-determination, as well as mutual respect and understanding. Consequently, person-centred practice is defined as providing care that is responsive to the individuals’ preferences, needs and values, and which promotes active involvement in their own care[3]. Nurses and midwives have significant roles in contributing to positive experiences of care. Measures that evaluate the impact of care, using a person-centred approach, are important. Despite this, evidence demonstrates that greater emphasis continues to be placed on quantifiable measures and performance indicators.

An established research programme has previously developed a set of eight, person-centred nursing and midwifery key performance indicators (KPIs) (See Table 1). The KPIs differ from commonly cited quality indicators as they relate directly to key elements of person-centredness, and have potential to contribute to improving the care experience[4]. The KPIs have a strong theoretical basis, being underpinned by the Person-centred Nursing Framework[4] (See Figure 1). The measurement framework developed alongside the KPIs includes four methods to collect data to evidence the KPIs (See Table 2). These data collection methods all relate to one or more of the eight KPIs, with each KPI having at least two sources of data to inform it. For example, KPI 5 (*Time spent by nurses with the patient*) is informed by survey data, observations, and the patient stories. The KPIs and measurement framework has been evaluated in a range of contexts[5–7]. This work has been underpinned by the MRC guidelines for the development, evaluation and implementation of complex interventions[8]. Evaluation outcomes illustrate that the KPIs are: an effective measure to evidence performance of nursing teams; a powerful driver for improvements in practice; and a mechanism that can promote person-centred cultures[9]. For example, within acute paediatric inpatient settings, use of the KPIs was found to generate data that encouraged more effective communication with children and their families, and that prompted critical appraisals of practice associated with making changes in practice to improve patient experience[5]. Following evaluation of the paper-based version of the KPIs and measurement framework, further work examined a prototype, app-based version. Evidence indicated that even as a prototype, the app helped to make KPI data more accessible, capturing it in real-time and enabling it to be used more easily to improve the experience of care[10]. This reflects the importance of digital health technologies which represent the most realistic option for the collection of large-scale data pertaining to person-centred measures[11]. In particular, mHealth apps are considered to provide a feasible approach to the collection of this type of data, as they can be used at scale and at relatively low cost[11,12].

**Figure 1.**
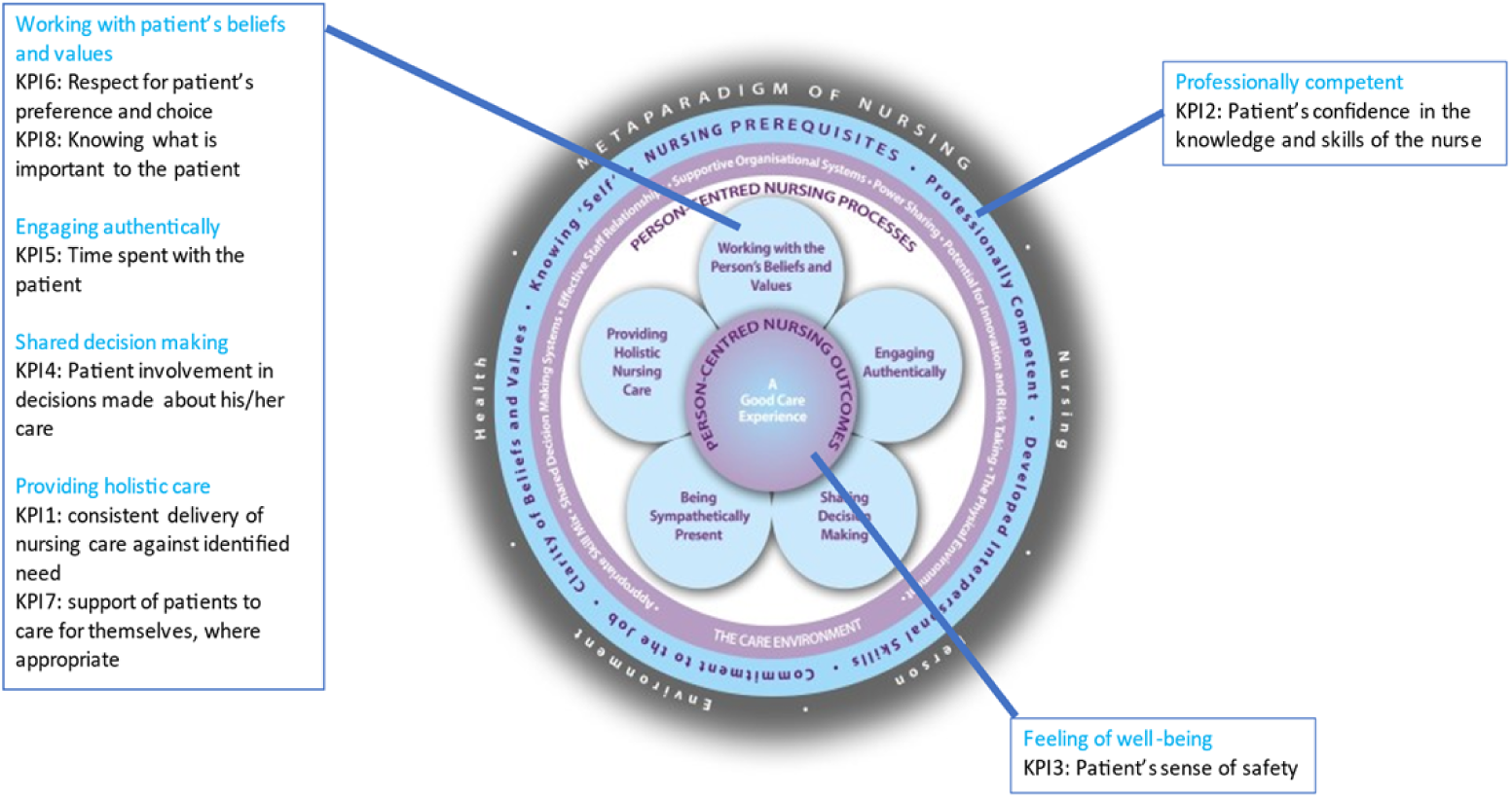
Key Performance Indicators mapped onto Person-centred Nursing Framework

**Table 1.**
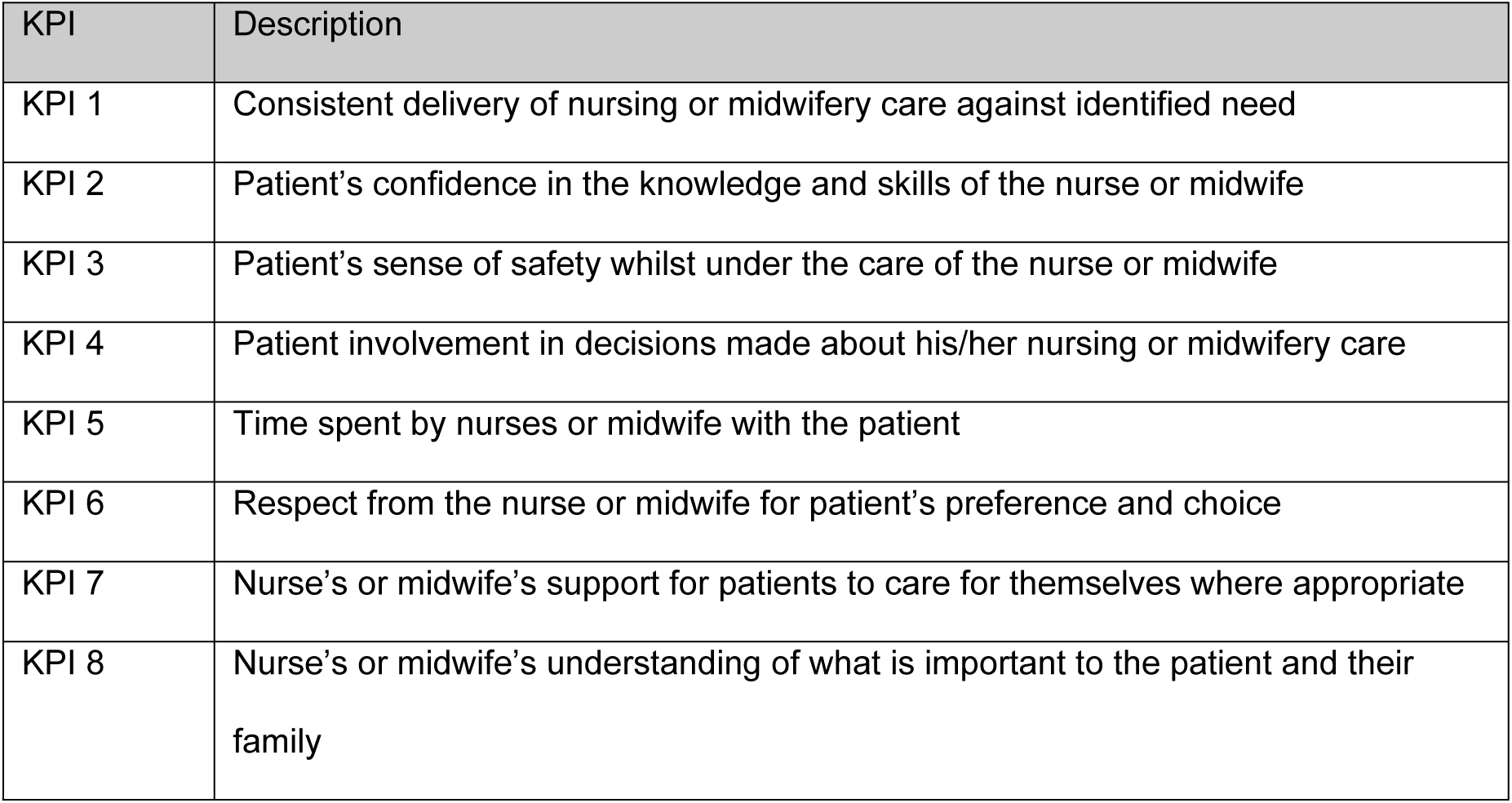
Key Performance Indicators.

| KPI | Description |
| --- | --- |
| KPI 1 | Consistent delivery of nursing or midwifery care against identified need |
| KPI 2 | Patient's confidence in the knowledge and skills of the nurse or midwife |
| KPI 3 | Patient's sense of safety whilst under the care of the nurse or midwife |
| KPI 4 | Patient involvement in decisions made about his/her nursing or midwifery care |
| KPI 5 | Time spent by nurses or midwife with the patient |
| KPI 6 | Respect from the nurse or midwife for patient's preference and choice |
| KPI 7 | Nurse's or midwife's support for patients to care for themselves where appropriate |
| KPI 8 | Nurse's or midwife's understanding of what is important to the patient and their family |

**Table 2.** The measurement framework for the iMPAKT App.

| Measurement tool | Description of tool | Key Performance Indicators assessed |
| --- | --- | --- |
| 1: Surveys for patients, carer, and partners | Administered via a questionnaire with Likert scale responses to eight questions relating to each KPI. | KPI 1-8 |
| 2: Patient Stories | Audio recording of patient speaking about their own experience of care. Aimed at capturing what is important to the patient, and what would improve their experience. Stories are then reviewed, and any relevant text related to any of the KPIs is “tagged” as “POSITIVE” or “NEGATIVE”. | KPI 1-8 |
| 3: Document Reviews | To establish if there is consistency between staff views and what is recorded in the patient documents for KPI 1 (Consistent delivery of care against identified need) and KPI 8 (Understanding of what is important to the patient). | KPI 1 and KPI 8 |
| 4: Activity Reviews / Observations of practice* | To capture the activity of staff (time spent with patients) during a single shift or when directly observing patient areas over 30 minutes. | KPI 5 |

\* The iMPAKT App can be accessed as a “community” version where only the “Activity Review” section is viewed, or can be accessed as a “Acute / Inpatient care” version, where the 30 minute “Observation” section is viewed.

Despite the potential usefulness of the iMPAKT App for the collection of person-centred measures, there are often organisational barriers to the use of digital tools in healthcare settings[13]. Well-designed implementation strategies are particularly important for complex, multilevel digital interventions[14]. These strategies are intended to facilitate the integration of an intervention within routine practice. These approaches can include efforts to guide translation into clinical practice, to understand determinants of implementation, or to evaluate the actual effectiveness of implementation strategies. While there is growing evidence focused on development of theories and frameworks to optimise intervention implementation[15,16], studies directly exploring implementation success have received limited attention to date. Furthermore, selecting appropriate strategies for implementing any mHealth intervention to enhance its acceptance and long-term sustainability can be challenging. Tailoring implementation can ensure that specific contextual considerations and needs are addressed, and that the most effective available strategies are used, dependent on the setting[17]. This is important and highlights the need to understand the factors that affect adoption and maintenance when programs are used in heterogenous settings[18]. Moreover, in order to successfully implement and integrate digital technologies in complex healthcare environments, different factors at the individual, organizational, and policy levels may need to be considered[19]. Some factors that are key to effective implementation are also not yet fully understood. This includes the impact on implementation from the perspectives of all levels of healthcare staff[20]. For example, it has been shown that where staff involved in facilitating quality improvement interventions have insufficient training and support at an organisational level, this can be associated with poorer implementation outcomes[21].

Following evaluation of the initial prototype app[10], a fully functioning version (the iMPAKT App) has been built using cross platform development. The core content and structure of the iMPAKT App is closely associated with the work which developed and evaluated the person-centred KPIs for nursing and midwifery[6–9]. The app includes sections that relate to the four measurement tools used for assessing each of the KPIs (See Table 2 and Figure 2). Data are collected using the app over a recommended period, or “Cycle”, of six weeks. Within each measurement cycle, a minimum data set is required that consists of 20 surveys, 3 patient stories, 10 document reviews and 3 assessments of time spent with patients. In addition to the four main sections of the app, a “Cycles” section, where new, six-week data collection periods can be started is included. A “Reports” section is also accessible for users to access previously collected data, allowing for comparisons to be made between different cycles. An “About the app” page provides information on the purpose and background of the app (See Figure 3 for selected screen shots).

**Figure 2.**
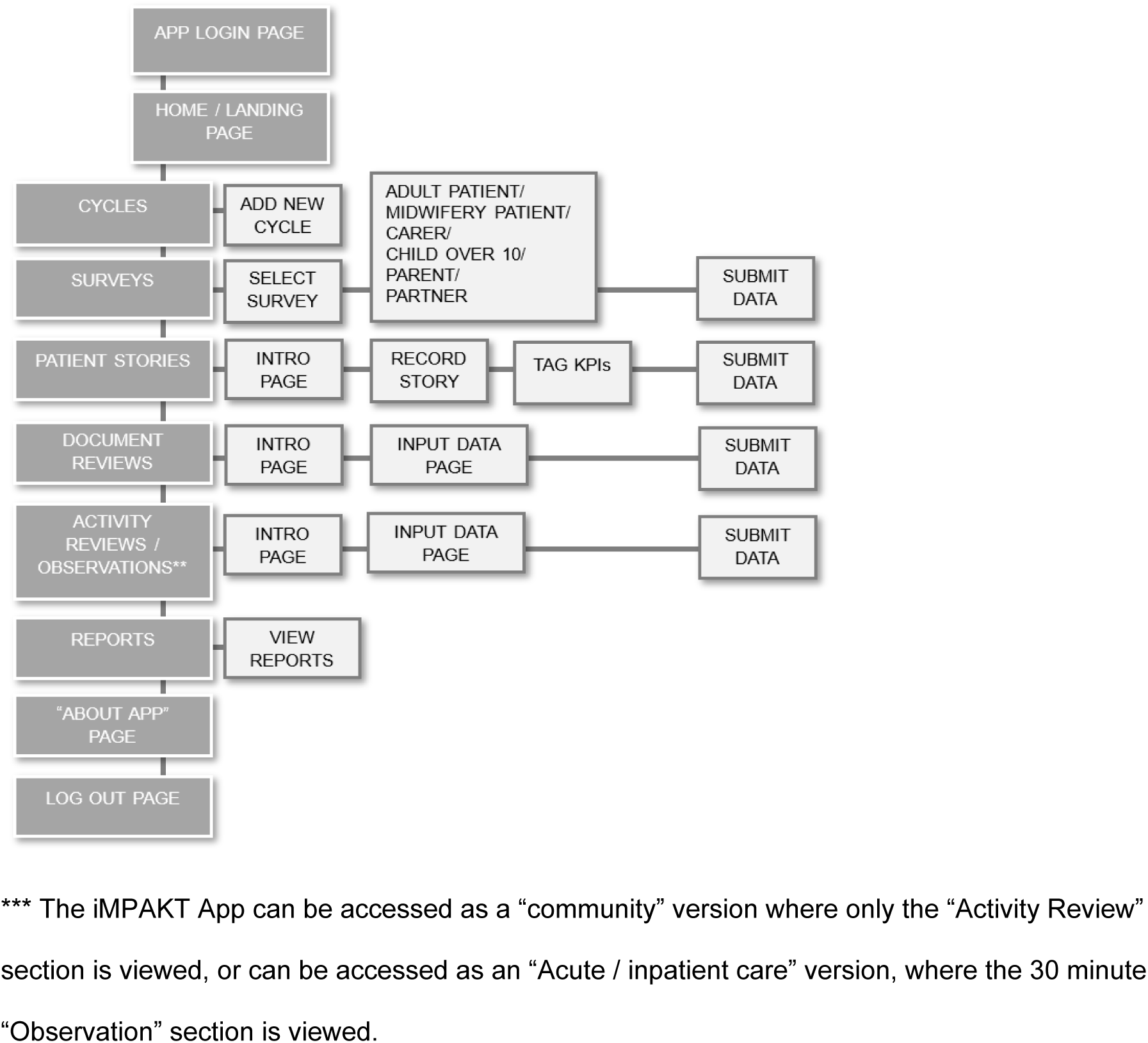
**Wireframe showing main sections of iMPAKT App**

**Figure 3.**
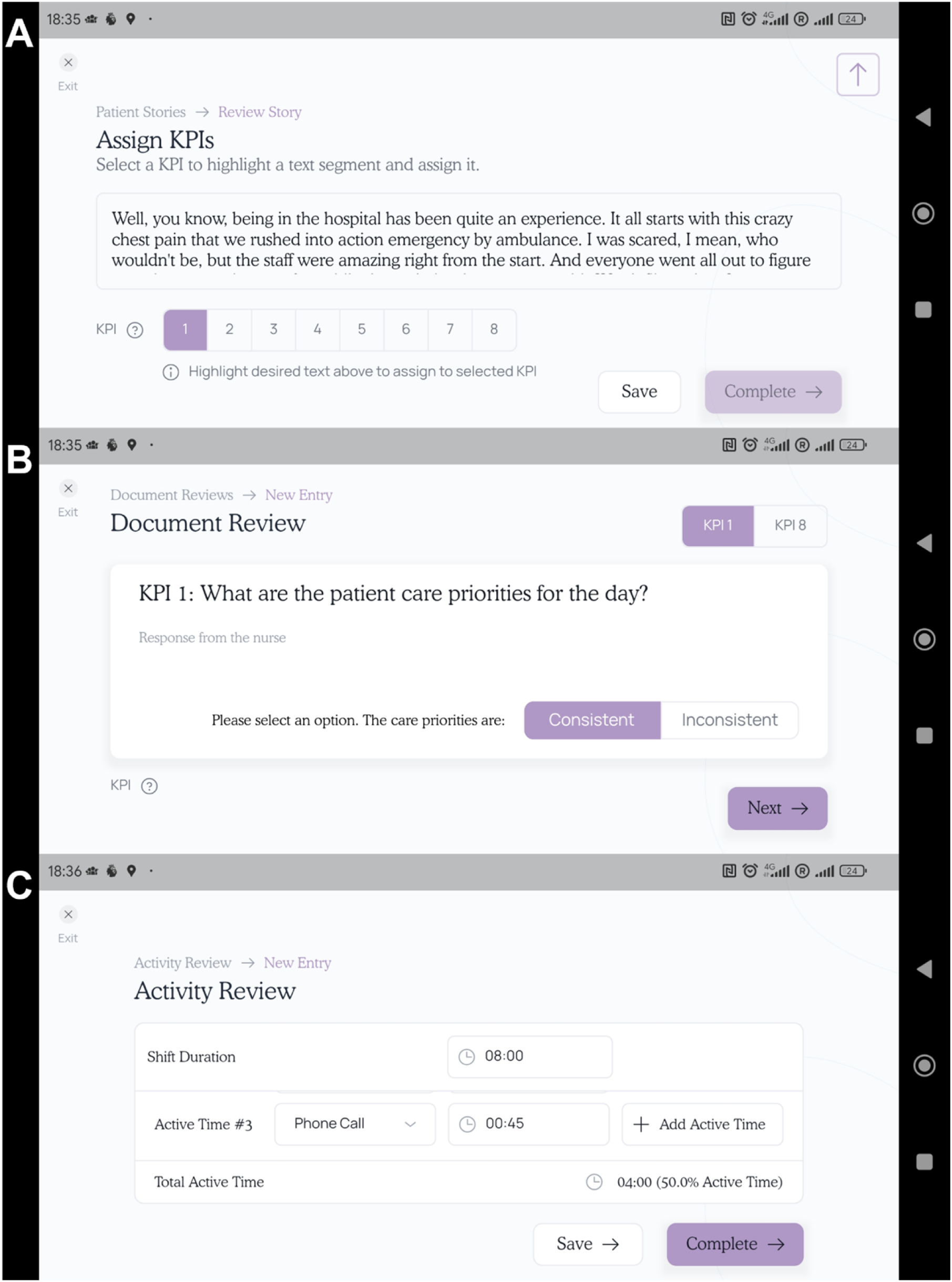
Selected screen shots from the revised version of the iMPAKT App showing pages from: A. the “Patient Stories”, B: the “Document Reviews”, and C. the “Activity Reviews” sections of the app

### Aim

The aim of this study is to evaluate the implementation of the iMPAKT App, a digital tool for implementing and measuring person-centredness in nursing and midwifery practice.

### Objectives

Specific objectives of the study are:

i. to examine acceptability and engagement with the app by nurses and midwives who use the app during the evaluation period.
ii. to explore user perspectives on factors that influence implementation of the app across different practice settings and contexts.
iii. to explore the potential for the app to effectively record large-scale data on person-centred measures that can be used to evaluate the impact of initiatives designed to improve person-centredness in nursing and midwifery practice.

Results will be used to develop an understanding of the determinants and processes underpinning successful implementation of the app. These findings will help to inform further research to develop tailored implementation strategies, and facilitate wider implementation and large scale collection of data on person-centred measures using the iMPAKT App.

## METHODS

### Theories and frameworks guiding the iMPAKT App implementation study

Various methods exist for applying implementation strategies, including taxonomies and the use of systematic approaches[22]. Theories and frameworks such as the Consolidated Framework for Implementation Research (CFIR)[23] offer a flexible approach, incorporating multiple factors at the intervention, individual, organizational, and environmental levels. The CFIR includes five domains (intervention characteristics, outer setting, inner setting, characteristics of individuals and process), with 39 underlying constructs that may influence implementation. The framework provides a practical guide for implementors to systematically assess multi-level potential barriers and facilitators to implementation during the planning, execution and evaluation of interventions. The study will follow the Standards for Reporting Implementation Studies (StaRI) recommendations[24].

### Study design and setting

This implementation study will use a convergent, multiple-methods approach to evaluate implementation of the iMPAKT App across different contexts and settings. The implementation study will last for 16 months in total and the implementation process will involve completion of two separate data cycles using the app (each lasting 6 weeks), with a 4 weeks interval in between cycles. Seven organisations (study sites) from the United Kingdom and Australia will be included in the study. These organisations represent a range of acute / inpatient, and community care settings. During the study, nursing and midwifery teams from within the study sites will be recruited to take part in implementation testing of the IMPAKT App. Initial approaches to service and team leads from each study site will be made to explore organisational readiness to participate in the study; and to identify teams suitable for inclusion in the study. Purposive sampling will be used to recruit individual participants from a range of teams that are selected based on their area of practice and team size. e.g., acute care wards, district nursing, or community-based, teams. The sites will begin the study asynchronously, with teams from each site recruited at different timepoints within the 16 month study period. The study will aim to recruit at least one, but no more than five teams from each site. A maximum number of 20 teams will be recruited to ensure that data can be collected and fully analysed during the study evaluation period.

During the study, participants will collect data on person-centred measures using the iMPAKT App. This will be done using the four measurement tools in the app as part of usual care procedures to collect data on experience of care. The following, non-directed strategies will be used to support implementation of the app. Firstly, all participants will be asked to take part in a two week preparation phase which will include training and app familiarisation. As part of this phase, participants will be asked to attend a two hour workshop (delivered in person or online, based on preference). Participants will also be provided with a weblink containing written information and a demonstration video on how to use the iMPAKT App. Members of the research group will also attend meetings with the teams at the study sites, the frequency of which will be based on the preference of the teams. The content of the meetings will include any technical support needed for teams using the app. Each team will be asked to identify at least one iMPAKT “Champion”, who will help to support implementation of the app.

As part of the familiarisation process, posters will also be placed in clinical areas during the implementation study in order to promote awareness of the app.

Prior to data collection, the iMPAKT App will be downloaded onto touch screen devices (Apple Corporation, iPad 6^th^ Gen, or a more recent model). These are devices that are used routinely in the practice settings. Members of the team will be provided with login details for the app (staff email address and a four digit PIN which can be reset by the user). In this way, individual participants can record data using the app (using their own login details), but data can be collated and summarised for the whole team (meaning all team members will be able to see progress towards completion of the minimal data set for the current data cycle). During each data collection cycle, teams will be asked to record at least 20 surveys, 3 Patient Stories, 10 Document Reviews and 3 Assessments of time spent with patients. Each team will complete two separate data cycles using the app (each lasting 6 weeks), with both data collection cycles being separated by a 4 week interval, during which data from the first cycle can be reviewed by the team. This will mean that each team will participate in the study for approximately 4 months.

### Study participants and sample size

Participants in the study will be: 1. Nurses or midwifes from the clinical teams who are recruited to the study. 2. Team / service leads or deputies. An accurate sample size is difficult to establish given the expected variation in the size and number of teams recruited at each site. However, given the planned number of teams (between 7 and 14), it is anticipated that the number of individuals recruited will provide an adequate samples size for this type of study design[25]. After service or team leaders have identified and approached the teams who take part in the study, all members of the teams will be invited by a researcher (in person, or via email) to take part in the evaluation. They will be provided with a participant information sheet and given a minimum of 48 hours to consider the study information and ask any questions. If willing to take part, they will then be asked to provide written, informed consent to take part in the evaluation. Not all team members will be required to take part in the evaluation if they do not wish to do so. Where one or more team members do not consent to take part, the team may still be recruited to the study.

### Data collection

Qualitative and quantitative implementation data will be collected during the study evaluation period, as described below. Qualitative methods will be used to explore individual responses, views and experiences around acceptability and engagement with app, and its implementation, as well as to examine variations in contexts[26]. Individual or small group (n=2-3) semi-structured interviews and focus groups (n=4-8) will be conducted face-to-face at study sites, or remotely via teleconference. Although there is no set rule about focus group size, it is suggested that small group interviews, or “mini focus groups” (made up of two to four members), are suitable in situations where participants have specialized knowledge and/or experience to discuss a subject[27]. If teams are large in number, or where there are multiple teams within a site, focus groups may be used, but this choice will be based on individual or team preference. All interviews and focus groups will take place following completion of the second cycle of data collection using the app (Week 18), and will be conducted by two experienced qualitative researchers (TMcC; SOC). These will be guided by questions derived from the Consolidated Framework for Implementation Research (https://cfirguide.org/) (See Appendix 5 for interview and focus group schedule). A flexible approach will be used when carrying out the interviews based on the CFIR constructs. Not all questions from the schedule will be used during interviews but the schedule provides a structure to guide any discussions around implementation issues that arise during interviews or focus groups. The qualitative component of the study will follow the consolidated criteria for reporting qualitative research (COREQ) criteria[28] All interviews and focus groups will be audio-recorded and transcribed verbatim, and will be expected to last between 45 and 90 minutes.

Quantitative data will be recorded at the end of each cycle of data collection (Week 8 and Week 18) (See Figure 4). This will include: 1. The number of person-centred measures (surveys, patient stories, record reviews and observations of practice) that were recorded by each team during the data collection cycle. 2. App usage statistics (time spent using the app, number of app logins). 3. Completion by each participant of the System Usability Scale (SUS)[29,30]. This is a 10-item measure of digital intervention usability. Items are scored on a 5-point Likert scale (Strongly Disagree-Strongly Agree) with an overall score calculated out of 100. A minimum of 10 direct observations of participants using the iMPAKT App will be made at each study site. Observations will be recorded using a semi-structured observation tool consisting of a 4-point rating scale, asking the following question: “Was implementation as intended”. The scale will range from 0 indicating “No/ Very low” implementation, to 4, indicating “Very high” implementation. This tool will be used to assess each component of the iMPAKT App. Open text boxes will be used to document further observations. These findings will assess fidelity and any contextual factors required to facilitate implementation of the app.

**Figure 4.**
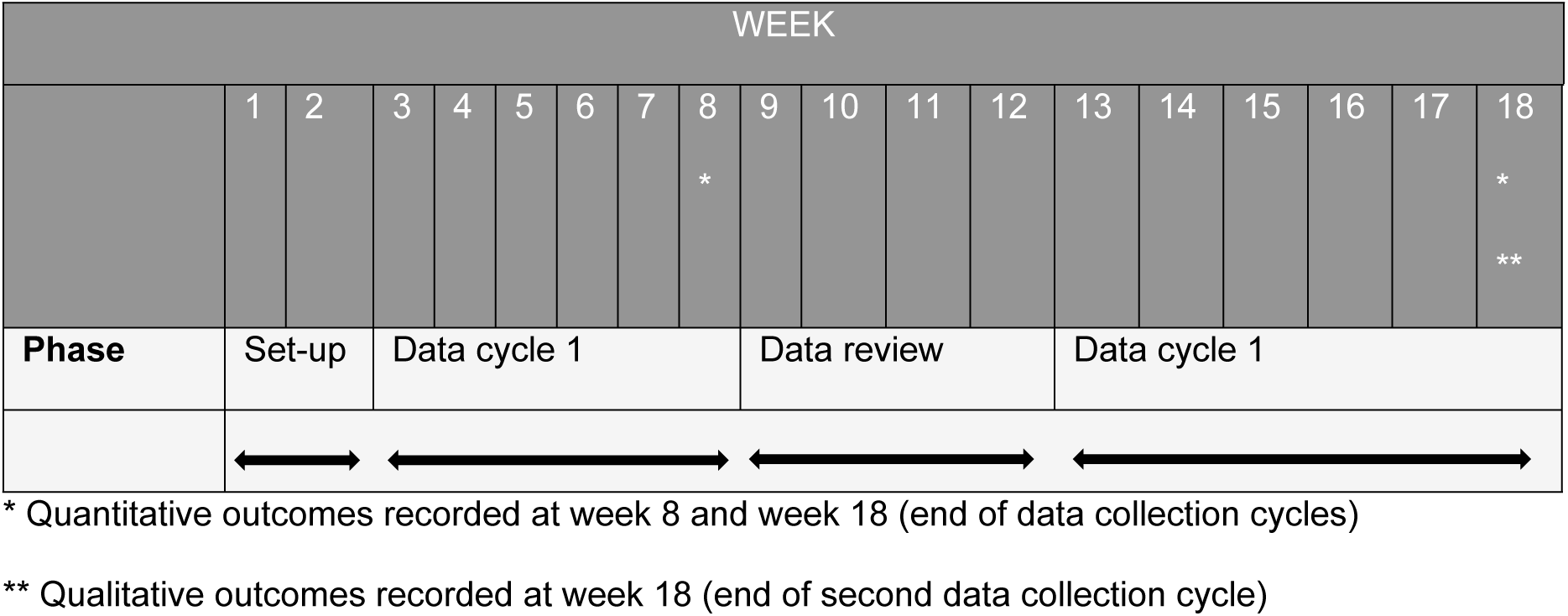
**Time spent in each phase by teams recruited to the study**

### Data analysis

For all qualitative data, a directed content analysis approach based on deductive and inductive coding will be used[26]. An initial coding scheme will be developed. Following line-by-line coding, findings will be summarised based on the coding scheme and used to revise initial codes if necessary and develop new codes based on emerging data. Respondent validation will be undertaken for approximately 10% of the interviews. Related codes will then be clustered and grouped into initial themes based on CFIR constructs. Narrative summaries will be written for each theme, then reviewed and discussed to refine main themes and sub-themes to ensure coherence. NVivo version 12 will be used to manage data and facilitate the analysis process, which in summary included the following stages: i. Independent transcription, ii. Data familiarisation, iii. Independent coding, iv. Development of an analytical framework, v. Indexing, vi. Charting and vii. Interpreting data.

For the quantitative data analysis, all data recorded during the two data collection cycles will be downloaded as excel files for each team. The SUS will be scored /100 using an online excel form (www.measuringux.com/SUS_Calculation.xls). Descriptive statistical analysis will be used to analyse quantitative data. Responses and measurements from all data sources will be presented as numbers and percentages for categorical variables and mean and standard deviation or median and interquartile intervals for continuous variables. This will help to understand the level of satisfaction and perceived utility associated with implementation of the app, and explain the extent to which it is used as intended. The quantitative data will be compared between time-points using Chi-squared test or Fisher’s exact tests to compare categorical variables. Independent t-test or Wilcoxon rank-sum test will be used to compare continuous variables.

### Ethics

Ethical approval for the study Approval for the study was granted by the Nursing and Health Research Ethics Filter Committee at Ulster University (Reference: FCNUR-24-011). No participants will be enrolled into the study prior to review by relevant regulatory authorities and REC approval. Participants in the evaluation will be provided with a Participant Information Sheet and Consent Form. Written, informed consent will be obtained from all participants and only de-identified data will be analysed and reported. No personal or identifiable patient /carer or supporter information will be recorded by the app.

### Patient and Public Involvement

Members of an established PPI group have been consulted on all aspects of the iMPAKT project, including design and development of the app, user testing of the app, and co-design of the evidence informed implementation strategy for the app.

## DISCUSSION

This protocol outlines the systematic methods for the evaluation of a complex quality improvement intervention based on using the iMPAKT App to improve and measure person-centredness in nursing and midwifery care. The evaluation will allow for greater understanding of the impact of the intervention and the context in which this impact occurs. It will assess successes and failures related to implementation, in addition to determining factors associated with scale-up and adaptation for other care settings. The study will contribute to stronger evidence around use of quality improvement interventions, and efforts to develop evaluation methods in digital health studies. The study will also assess the potential for large scale collection of data on person-centred care that can contribute to addressing challenges within current healthcare systems occurring as a result of the changing age profile of the population, and increases in the numbers of people living with long-term, multiple health conditions. A key strength of the proposed study is the use of multiple-methods, which can contribute to a more comprehensive understanding of implementation. This approach will provide a detailed analysis based on the perspectives of a range of participants working in different contexts and settings, and will provide in-depth knowledge on utility, barriers, and adoption. Findings from this evaluation will also provide valuable insights on wider adoption and scaling-up of the implementation strategies used.

## Data Availability

Deidentified research data will be made publicly available when the study is completed and published.

## Acknowledgments

We acknowledge the valuable contributions of the app development team at Miroma Project Factory (Australia), the iMPAKT Project Advisory Board, and the study teams at University College London Hospitals NHS Foundation Trust, NHS Lothian, the Northern Health and Social Care Trust, Northern Ireland, the South Eastern Health and Social Care Trust, Northern Ireland, the Northern Adelaide Local Health Network (NALHN) and Prince of Wales Hospital, Sydney.

## Author Contributions

**Conceptualization:** Sean O’Connor, Donna Brown, Ian Cleland, Valerie Wilson, Tanya McCance.

**Methodology:** Sean O’Connor, Donna Brown, Ian Cleland, Valerie Wilson, Tanya McCance.

**Writing – original draft:** Sean O’Connor, Tanya McCance.

**Writing – review & editing:** Sean O’Connor, Donna Brown, Ian Cleland, Valerie Wilson, Tanya McCance.

## Data Availability

Data is available on request from the corresponding author.

## Appendix 1 Semi-structured interview and focus group schedule

The following questions and prompts are based on the Consolidated Framework for Implementation Research (https://cfirguide.org/). Questions should not necessarily be used or worded in exactly the same way, as some flexibility may be required in different contexts.

### Intervention Characteristics

#### Evidence Strength & Quality

1. What kind of information are you aware of that shows whether or not the iMPAKT App works in your setting?

PROMPT: How could this knowledge affect your perception of the intervention?

2. What do you think influential stakeholders would think of the iMPAKT App?

3. What kind of supporting evidence about the effectiveness of the iMPAKT App would be needed to get staff on board?

#### Relative Advantage

4. How does the iMPAKT App compare to other similar approaches you have seen in your setting?

PROMPT: What advantages/disadvantages does the intervention have?

#### Adaptability

What kinds of changes could make the iMPAKT App work most effectively in your setting?

PROMPT: Do you think you will be able to make these changes? Why or why not?

5. Are there components that should not be altered?

#### Trialability

6. Do you think it would be possible to pilot the iMPAKT App before making it available to everyone?

PROMPT: Why or why not?

#### Complexity

7. How complicated is it to implement the iMPAKT App?

PROMPT: Consider the duration, scope, intricacy and number of steps involved and whether the app reflects a departure from previous practices.

#### Design Quality & Packaging

8. What support, such as online resources, or a toolkit, could help implement the iMPAKT App?

PROMPT: How should these be accessed?

9. How will available materials affect implementation of the iMPAKT App in your setting?

#### Cost

10. What costs will be incurred to implement the iMPAKT App?

### Outer Setting

#### Patient Needs & Resources

11. How well do you think the iMPAKT App will meet the needs of the individuals served by your organization?

PROMPT: In what ways will the intervention meet their needs?

12. How do you think the individuals served by your organization will respond to the iMPAKT App?

#### Peer Pressure

13. To what extent would implementing the iMPAKT App provide an advantage for your organization compared to other organizations in your area?

#### External Policies & Incentives

14. What kind of local or national performance measures, policies, or guidelines influence implementation of the iMPAKT App?

### Inner Setting

#### Structural Characteristics

15. How could infrastructure of your organization (social architecture, age, maturity, size, or physical layout) affect the implementation of the iMPAKT App?

16. What kinds of infrastructure changes will be needed to accommodate the iMPAKT App?

PROMPT: Changes in scope of practice? Changes in formal policies? Changes in information system?

PROMPT: What kind of approvals will be needed? Who will need to be involved? PROMPT: Can you describe the process that will be needed to make these changes?

#### Networks & Communications

17. Are meetings, such as staff meetings, held regularly?

PROMPT: Who typically attends?

PROMPT: What is a typical agenda? How helpful are these meetings?

18. How do you typically find out about new information, such as new initiatives, accomplishments, issues, new staff, staff departures?

#### Culture

19. How do you think your organization’s culture (general beliefs, values, assumptions that people embrace) affect implementation of the iMPAKT App?

PROMPT: Can you describe an example that highlights this?

#### Implementation Climate

20. What is the general level of receptivity in your organization to implementing the iMPAKT App?

#### Tension for Change

21. How essential is the iMPAKT App for meeting the needs of the individuals served by your organization or other organizational goals and objectives?

#### Compatibility

22. How well does the intervention fit with existing work processes and practices in your setting?

PROMPT: What are likely issues or complications that may arise?

#### Relative Priority

23. What is the priority for iMPAKT App implementation relative to other initiatives that are happening now?

#### Organizational Incentives & Rewards

24. What kinds of incentives are there to help ensure that the implementation of the intervention is successful?

#### Goals & Feedback

25. Have you/your unit/your organization set goals related to the implementation of the iMPAKT App?

PROMPT: [If yes] What are the goals?

#### Leadership Engagement

26. What kind of support or actions can you expect from leaders in your organization to help make implementation successful?

#### Available Resources

27. Do you expect to have sufficient resources to implement and administer the intervention?

PROMPT: [If Yes] What resources are you counting on? Are there any other resources that you received, or would have liked to receive?

PROMPT: What resources will be easy to procure? [If no] What resources will not be available?

#### Access to Knowledge & Information

28. Who do you ask if you have questions about the intervention or its implementation?

PROMPT: How available are these individuals?

### Characteristics of Individuals

#### Knowledge & Beliefs about the Intervention

29. Do you think the iMPAKT App could be successfully implemented in your setting?

PROMPT: Why or why not?

#### Self-efficacy

30. How confident are you that you will be able to successfully implement the intervention?

PROMPT: What gives you that level of confidence (or lack of confidence)?

31. How confident do you think your colleagues feel about implementing the intervention?

PROMPT: What gives them that level of confidence (or lack of confidence)?

### Process

#### Champions

32. Other than the formal implementation leader, are there people in your organization who are likely to champion (go above and beyond what might be expected) the intervention?

#### Key Stakeholders

1. 33. What steps have been taken to encourage individuals to commit to using the intervention?
2. 34. What is your communication or education strategy (not including training, see Access to Knowledge and Information) for getting the word out about the intervention?

#### Reflecting & Evaluating

35. What kind of information could be collected when implementing the iMPAKT App?

PROMPT: Which measures? How will they be tracked?

PROMPT: How could this information be used?

36. How will you assess progress towards implementation or intervention goals?

## Appendix 2 System Usability Scale

System Usability Scale
|  |  | Strongly<br>agree |  |  |  | Strongly<br>disagree |
| --- | --- | --- | --- | --- | --- | --- |
| 1 | I would like to use the iMPAKT app frequently | 1 | 2 | 3 | 4 | 5 |
| 2 | I found the iMPAKT app to be complicated | 1 | 2 | 3 | 4 | 5 |
| 3 | I thought the iMPAKT app was easy to use | 1 | 2 | 3 | 4 | 5 |
| 4 | I think that I would need the support of a technical person to be able to use the iMPAKT app | 1 | 2 | 3 | 4 | 5 |
| 5 | I found the various functions in the iMPAKT app were well integrated | 1 | 2 | 3 | 4 | 5 |
| 6 | I thought there was too much inconsistency in the iMPAKT app | 1 | 2 | 3 | 4 | 5 |
| 7 | I would imagine most people would learn to use the iMPAKT app very quickly | 1 | 2 | 3 | 4 | 5 |
| 8 | I found the iMPAKT app very cumbersome to use | 1 | 2 | 3 | 4 | 5 |
| 9 | I felt very confident using the iMPAKT app | 1 | 2 | 3 | 4 | 5 |
| 10 | I needed to learn a lot before I could get going with the iMPAKT app | 1 | 2 | 3 | 4 | 5 |

© Digital Equipment Corporation, 1986

